# The association between periodontitis and the impact of oral health on the quality of life of individuals with psoriasis and psoriatic arthritis

**DOI:** 10.1101/2024.03.13.24304243

**Authors:** Amanda Almeida Costa, Luís Otávio Miranda Cota, Rafael Paschoal Esteves Lima, Alcione Maria Soares Dutra Oliveira, Sheila Cavalca Cortelli, José Roberto Cortelli, Renata Magalhães Cyrino, Victor Silva Mendes, Tarcília Aparecida Silva, Fernando Oliveira Costa

## Abstract

**Objective:** To evaluate the association between psoriasis (PSO), psoriatic arthritis (PsA) and periodontitis (PE), and the Oral Health-Related Quality of Life (OHRQoL) impacts on individuals with psoriatic disease’s daily activities compared to the non-psoriatic ones.

**Materials & Methods:** 296 individuals with psoriatic disease (PSO n= 210, APS n= 86) (cases) and 359 without these diseases (controls) were included. Complete periodontal examinations and collection of variables of interest were performed. The Brazilian version of the *Oral Impacts on Daily Performance* (OIDP) instrument was applied.

**Results:** The prevalence of PE was higher in PsA (57.0%; OR=2.67 95%CI 1.65–4.32; p<0.001) than in PSO (34.3%; OR=1.05 95% CI 0.73–1.51; p<0.001) compared to controls (33.1%). Both PsA and PSO groups showed more sites and teeth with 4-6mm probing depth (PD) and had higher OIDP scores than controls (p<0.001), thus indicating worse self-reported quality of life. PE, PSO+PE and consumption of alcohol/anxiolytics significantly influenced OHRQoL (p<0.05). The influence of periodontal parameters on OHRQoL was observed for the presence of PE; PD >6 mm; clinical attachment level >6 mm; higher plaque index, % sites and teeth with bleeding on probing (p<0.05).

**Conclusion:** Negative impacts of PE on the OHRQoL were demonstrated. The ones having PSO and especially PsA and PE presented significantly worse indicators.

## Introduction

The term periodontal diseases encompass changes that affect periodontal tissues which protect and insert the teeth, such as gingivitis and periodontitis (PE). PE is characterized by host-mediated, microbiologically associated inflammation, resulting in loss of periodontal attachment(1). Susceptibility to PE is related to polymicrobial disorders that result in dysbiosis considering its complex pathogenesis. The precise mechanisms that lead to the onset of PE are not completely known yet, although studies have shown that PE onset and progression depend on an imbalance in the individual’s immunoinflammatory response to dental biofilm microorganisms. Since the presence of biofilm is essential but not sufficient for PE onset, the recognition of microbial components by defense cells and the production of inflammatory mediators is indeed crucial(2). This recognition can activate the acquired immune response, leading to the differentiation of T-helper cells (Th) into Th1, Th2, Th17 or T-regulatory cells (Treg)(3). Th1 and Th2 cells of the adaptive immune system are the main agents in the pathogenesis of immune-mediated diseases. Th17 cells and the cytokines IL-17 and IL-23 seem to play fundamental roles(3).

Since the American Academy of Periodontology (1999) workshop considerable evidence has emerged regarding potential effects of PE on systemic diseases such as endocrine, cardiac, autoimmune disorders and adverse pregnancy outcomes(4–6).

Psoriasis (PSO) is a chronic inflammatory skin disease characterized by the inflammation of the dermis and epidermis. Its prevalence varies due to country-specific variations and the methodological variability of epidemiological studies, but in a global analysis, it has been outlined to vary from 0.14% to almost 3%. Given the growing and the aging of worldwide population and the fact that PSO primarily affects the adults, the burden associated with PSO may continue to increase(7). Psoriatic arthritis (PsA) is a chronic inflammatory musculoskeletal disease that has been defined as an inflammatory arthritis associated with PSO and seronegative for rheumatoid factor. PsA occurs in 2-3% of the general population, but among patients with PSO the prevalence varies from 6% to 42%, with involvement of the skin, nails, bones, tendons, ligaments, synovial membrane and joints(8,9).

Increasing evidence supports the association between PSO, PsA and multiple comorbidities including obesity, metabolic syndrome, cardiovascular disease, cerebrovascular disease, anxiety and depression (10,11) and PE (6,12–15).

The hypothesis of an association between these diseases is that PE can play a direct and indirect role in the development or exacerbation of psoriatic disease, as both consist of a complex interrelationship between cell types of the innate and acquired immune system, immunological factors produced by T cells, dendritic and Th1-mediated cells, through the IL-23/IL-17 axis and because they share some risk factors (e.g., diabetes and smoking) (16,17).

The relationship between PE, PSO and especially PsA has gaps and the pathophysiological paths of this association are not completely elucidated. To date, only 4 studies have been reported on the association of PE and PsA. (12–14,18) Hence, literature is significantly scarce in relation to studies that address the impact of OHRQoL of individuals with psoriatic disease.

This study is then justified by the need for greater knowledge about the potential association between PSO, PsA and PE, and the similarities in their comorbidities and risk factors, and also to seek a greater understanding of OHRQoL of the affected individuals. Thus, we hypothesized that higher OIPD scores are associated with the presence of worse periodontal clinical conditions in individuals with psoriatic disease.

The aim of this case-control study was to evaluate periodontal status, its clinical and epidemiological aspects and the impact of OHRQoL in PSO and PsA individuals compared to those without psoriatic disease.

## Materials and Methods

### Sampling

The sample size calculation was based on a variation of around 10% prevalence of PE among individuals with psoriatic disease, as reported by Mendes *et al*.(19) and Mishra *et al*.,(13) and a PE prevalence of 30-40% among controls and 50-60% among psoriatic individuals was assumed. The calculation was performed using the Fleiss method, with continuity correction using statistical software^3^. Based on a significance level of 0.05, 80% power and a case-control ratio of 1:2, 77 cases and 153 controls were determined to be the minimum required. However, during the data collection period (April 2020 – December 2022), 655 individuals were examined and considered eligible. The final sample consisted of 297 individuals in the case group, 210 individuals with PSO and 86 with PsA, and 359 without psoriasis (controls).

PSO individuals were selected from those undergoing treatment and monitored at the Department of Dermatology at Hospital Eduardo de Menezes, and at the Center for Specialized Medicine, Teaching and Research in Belo Horizonte, Brazil, from March 20, 2020 until July 30, 2022. The control group consisted of individuals without any dermatological disease, selected from relatives, companions or employees of the respective reference centers.

The following inclusion criteria were established: individuals aged 35 to 65 years, presence of **≥**14 teeth(20) and absence of contraindications for clinical periodontal examination. Exclusion criteria included individuals undergoing antibiotic therapy or periodontal treatment over the last three months. During clinical periodontal examinations, third molars were included only when in the position of second molars (19).

This study was approved by the Research Ethics Committee from the Federal University of Minas Gerais – Brazil (#CAAE 20156019.0.0000.5149) and followed the 1975 Declaration of Helsinki as revised in 2013. All participants were informed about the study procedures and signed a consent form before participating in the study.

### Variables of interest

The following variables of interest were collected during the interviews: gender, age, family income, educational level, flossing (yes/no), toothbrushing frequency (times/day), body mass index (BMI), diabetes(21), use of anxiolytics and antidepressants, smoking(22) and alcohol consumption (yes, no/occasional).

### Quality of Life assessment

A Brazilian version of the *Oral Impacts on Daily Performance* (OIDP) questionnaire was applied to assess OHRQoL (23,24). It assesses the frequency and severity of oral impact on individuals’ daily performances, as such: (1) physical performance (eating and enjoying food, speaking and hearing clearly, teeth cleaning); (2) psychological performance (sleeping and relaxing, smiling showing teeth without embarrassment, maintaining balanced emotional state); (3) social performance (throughout their main job or social role, enjoying contact with people). The occurrence of any difficulty is recorded dichotomously (yes/no) and the frequency is indicated by the duration period with the following scores: (1) lesser than once a month, or an interval of up to 5 days total; (2) once or twice a month, or an interval of up to 15 days total; (3) once or twice a week, or an interval of up to 30 days total; (4) three or four times a week, or an interval of up to 3 months total and (5) every or almost every day, or an interval of more than 3 months total. Regarding severity, it varies between “no severity” (0) and “extremely severe” (5). The maximum score is 200 and it is equivalent to multiplying the frequency by the severity, with the final score given by the sum of the difficulties reported in the 8 categories (200= 8 categories x 5 frequencies x 5 severity scores). The higher the score, the worse the OHRQoL is shown.

### Clinical periodontal examination

The level of oral hygiene was assessed using the plaque index (PI) (25). Scores for each site were added and mean values were recorded per tooth and individual. Periodontal parameters were then evaluated in all teeth, and values for each of the four sites (mesial, distal, buccal and lingual) were recorded: probing depth (PD), clinical attachment level (CAL), bleeding on probing (BOP). A manual periodontal probe model UNC-15, a clinical mirror and gauze were used.^4^

### Intra and inter-examiner agreement

Examinations, interviews and the OIDP questionnaire application were conducted by three experienced and trained periodontists (AAC, FOC and VSM). Intra-and inter-examiner agreements for periodontal examinations were performed in a pilot study with 12 individuals using the Kappa test and the intraclass correlation coefficient (ICC). The Kappa values for PD and CAL were >0.93 and the ICC >0.90.

### Definition and staging of periodontitis

Stages I, II, III and IV were included, according to the Tonetti *et al* (2018) criteria (1).

### Diagnosis of psoriasis and psoriatic arthritis

The diagnosis and severity of the disease (mild, moderate or advanced) were established by the group of dermatologists associated with the reference centers.

The psoriasis area and severity index (PASI)(26) was used and made available in all medical records. The PASI examines four regions of the body: (I) head and neck, (II) hands and arms, (III) chest, abdomen and back (trunk) and (IV) buttocks, thighs and legs. Each region is given a score to show how much each is affected by PSO (area) and another for how bad the PSO is (severity). The area score can range from 0 (no psoriasis) to 6 (all skin affected). Each region’s severity score is evaluated by adding redness, thickness, and scale scores, each of which is rated from 0 to 4, with a maximum of 12. The following cutoffs are used for severity: <7 mild, 7–12 moderate and >12 advanced psoriasis.

For the PsA diagnosis, The CASPAR criteria (27) was used: presence of confirmed inflammatory joint disease (joints, spine, or entheses); with at least three of these elements: current PSO, history of PSO or family history of PSO; dactylitis; bone formation in articular area (hands/feet); negative rheumatoid factor and psoriatic nail dystrophy. The sensitivity and specificity of the CASPAR criteria are 99.7% and 99.1%, respectively.

### Statistical analysis

Initially, the groups were compared in relation to the following variables: gender, age, family income, educational level, use of dental floss, BMI, diabetes, continuous use of anxiolytics, antidepressants, smoking and alcohol consumption using Chi-square, Fischer and Mann-Whitney tests when appropriate. Regarding periodontal parameters (IP, BOP, PD, CAL) the values per individual were obtained by summing the measurements from all periodontal sites and expressed as means and/or percentages.

An indicator was created in relation to OHRQoL, calculating the product between frequency and severity, multiplied by 100 and divided by 200. In the descriptive analysis of qualitative variables, absolute and relative frequencies were used, while for quantitative variables, measures of position, central tendency and dispersion were used. To compare categorical variables regarding QoL indicators, the Mann-Whitney and Kruskal-Wallis tests were used. The analysis of the frequency and severity of indicators of difficulty in carrying out daily activities using the OIDP are presented in Bootstrap intervals with a 95% confidence interval (CI). When the CI of an item does not overlap with one of another item within a group, there is statistical evidence that the items mean are statistically different. The software used in the analyzes was R.^5^ Results with p<0.05 were considered significant.

## Results

The sample consisted of 210 psoriasis individuals (PSO), 86 with PsA and 359 controls, 407 women and 248 men, with a mean age of 47.5 (±5.1) years for the control group, 49.7 (±4.2) years for the PSO group (p<0.001); and 48.8 (± 4.3) for the PsA group (p= 0.011) (Table 1). Regarding variables of interest, PSO individuals presented significantly higher values of the occurrence of diabetes (p=0.004), alcohol consumption (p=0.001), antidepressants (p=0.006) and anxiolytics (p = 0.008), as well as higher BMI (p <0.001) (Table 1) when compared to controls. PsA individuals also showed greater antidepressants (p<0.001) and anxiolytics use (p=0.013), diabetes (p=0.002) and higher BMI (p<0.001) compared to controls. In addition, data demonstrated PsA individuals had greater alcohol (p=0.040) and antidepressants (p=0.036) consumption than PSO individuals.

**Table 1.** Comparative analysis of variables of interest in the studied groups.

| Variables | Group |  |  | A-B |  | A-C |  | B-C |  |
| --- | --- | --- | --- | --- | --- | --- | --- | --- | --- |
|  | Control (A)<br>(n = 359) | Psoriasis<br>(B)<br>(n = 210) | Psoriatic<br>arthritis (C)<br>(n = 86) | p | OR<br>CI (95%) | p | OR<br>CI (95%) | p | OR<br>CI (95%) |
| **Gender** |  |  |  |  |  |  |  |  |  |
| Feminine | 225 (62.7%) | 128 (61.0%) | 48 (55.81%) | 0.683* | 1 | 0.984* | 1 | 0.768* | 1 |
| Masculine | 134 (37.3%) | 82 (39.0%) | 38 (44.19%) |  | 1.08 (0.76; 1.53) |  | 1.00 (0.61; 1.62) |  | 0.93 (0.55; 1.55) |
| **Age (years)** |  |  |  |  |  |  |  |  |  |
| Mean ± s.d. | 47.5 ± 5.1 | 49.7 ± 4.2 | 48.8 ± 4.3 | **<0.001**‡ | - | **0.011**‡ | - | 0.840‡ | - |
| **Income** |  |  |  |  |  |  |  |  |  |
| 1-2 Brazilian Minimum<br>Wage (BMW) | 146 (40.7%) | 84 (40.0%) | 33 (38.4%) | 0.439* | 1 | 0.926* | 1 | 0.636* | 1 |
| 3-4 BMW | 129 (35.9%) | 85 (40.5%) | 32 (37.2%) |  | 1.15 (0.78; 1.68) |  | 1.10 (0.64; 1.88) |  | 0.96 (0.54; 1.7) |
| 5 BMW or more | 84 (23.4%) | 41 (19.5%) | 21 (24.4%) |  | 0.85 (0.54; 1.34) |  | 1.11 (0.6; 2.03) |  | 1.3 (0.67; 2.53) |
| **Educational level** |  |  |  |  |  |  |  |  |  |
| < 8 years | 248 (69.1%) | 144 (68.6%) | 55 (64.0%) | 0.899* | 1 | 0.360* | 1 | 0.442* | 1 |
| > 8 years | 111 (30.9%) | 66 (31.4%) | 31 (36.0%) |  | 1.02 (0.71; 1.48) |  | 1.26 (0.77; 2.06) |  | 1.23 (0.73; 2.08) |
| **Smoking** |  |  |  |  |  |  |  |  |  |
| No | 314 (87.5%) | 183 (87.1%) | 69 (80.2%) | 0.911* | 1 | 0.082* | 1 | 0.129* | 1 |
| Yes | 45 (12.5%) | 27 (12.9%) | 17 (19.8%) |  | 1.03 (0.62; 1.72) |  | 1.72 (0.93; 3.18) |  | 1.67 (0.86; 3.25) |
| **Alcohol consumption** |  |  |  |  |  |  |  |  |  |
| No | 159 (44.3%) | 93 (44.3%) | 27 (31.4%) | 0.999* | 1 | 0.029* | 1 | **0.040** * | 1 |
| Yes | 200 (55.7%) | 117 (55.7%) | 59 (68.6%) |  | 1.00 (0.71; 1.41) |  | 1.74 (1.05; 2.87) |  | 1.74 (1.02; 2.95) |
| **Flossing** |  |  |  |  |  |  |  |  |  |
| No | 200 (56.0%) | 122 (58.1%) | 51 (59.3%) | 0.660* | 1 | 0.582* | 1 | 0.848* | 1 |
| Yes | 157 (44.0%) | 88 (41.9%) | 35 (40.7%) |  | 0.92 (0.65; 1.30) |  | 0.87 (0.54; 1.41) |  | 0.95 (0.57; 1.58) |
| **Brushing** |  |  |  |  |  |  |  |  |  |
| 1 time | 12 (3.3%) | 10 (4.8%) | 6 (7.0%) | 0.638* | 1 | 0.303* | 1 | 0.741† | 1 |
| 2 times | 104 (29.1%) | 63 (30.0%) | 25 (29.1%) |  | 0.73 (0.30; 1.78) |  | 0.48 (0.16; 1.41) |  | 0.66 (0.22; 2.01) |
| 3 or 4 times | 242 (67.6%) | 137 (65.2%) | 55 (64.9%) |  | 0.68 (0.29; 1.61) |  | 0.82 (0.59; 1.14) |  | 0.88 (0.59; 1.29) |
| **Antidepressant use** |  |  |  |  |  |  |  |  |  |
| No | 320 (89.9%) | 172 (81.9%) | 61 (70.9%) | **0.006** * | 1 | **<0.001** * | 1 | **0.036** * | 1 |
| Yes | 36 (10.1%) | 38 (18.1%) | 25 (29.1%) |  | 1.96 (1.20; 3.21) |  | 3.64 (2.04; 6.5) |  | 1.86 (1.04; 3.32) |
| **Anxiolytics use** |  |  |  |  |  |  |  |  |  |
| No | 306 (86.4%) | 161 (77.8%) | 65 (75.6%) | **0.008\*** | 1 | **0.013\*** | 1 | 0.684* | 1 |
| Yes | 48 (13.6%) | 46 (22.2%) | 21 (24.4%) |  | 1.82 (1.16; 2.85) |  | 2.06 (1.15; 3.67) |  | 1.13 (0.63; 2.04) |
| **Diabetes** |  |  |  |  |  |  |  |  |  |
| No | 333 (93.0%) | 180 (85.7%) | 71 (82.6%) | **0.004 \*** | 1 | **0.002 \*** | 1 | 0.492* | 1 |
| Yes | 25 (7.0%) | 30 (14.3%) | 15 (17.4%) |  | 2.22 (1.27; 3.89) |  | 2.81 (1.41; 5.61) |  | 1.27 (0.64; 2.5) |
| **Body mass index** |  |  |  |  |  |  |  |  |  |
| Mean ± s.d. | 24.8 ± 1.8 | 25.8 ± 2.2 | 26.6 ± 3.1 | **<0.001‡** | - | **<0.001‡** | - | 0.055‡ | - |
| **Quality of life** |  |  |  |  |  |  |  |  |  |
| Mean ± s.d. | 3.3 ± 6.4 | 12.6 ± 25.5 | 14.6 ± 27.0 | 0.052§ | - | **0.011 §** | - | 0.322‡ | - |
| **Quality of life** |  |  |  |  |  |  |  |  |  |
| Not impacted | 243 (67.7%) | 134 (63.8%) | 49 (57.0%) |  |  | 0.060* |  | 0.272* |  |
| Impacted | 116 (32.3%) | 76 (36.2%) | 37 (43.0%) | 0.345* | 1 |  | 1 |  | 1 |
|  |  |  |  |  | 1.19 (0.83; 1.70) |  | 1.58 (0.98; 2.56) |  | 1.33 (0.8; 2.22) |

Chi-square test (\*), Fisher's exact test (†), Student's t-test (‡) and Mann-Whitney test (§). Significant values in bold.

Table 2 shows the comparison between groups regarding periodontal parameters. PsA individuals showed significantly more PE (57.0%; OR=2.67, 95%CI: 1.65-4.32 p<0.001), while PSO (34.3%; OR=1.05; 0.73-1.51; p<0.001) than controls. PsA and PSO group revealed a greater number of sites and teeth with PD 4-6mm than controls (p<0.001). PD mean in PsA was significantly higher than in PSO (p=0.004) and controls (p=0.001), as well as in the comparison between PSO and controls (p<0.001). CAL frequency of >6mm was higher in PsA group, and significantly higher than in PSO group (p<0.001) and controls (p=0.001). Regarding BOP, higher mean values were observed in PsA, followed by PSO and controls (PsA>PSO>Controls; p<0.001) and (PSO>controls; p=0.034)]. As for IP; PsA group presented significantly higher and worse scores than those reported in the PSO group (p=0.001) and controls (p<0.001).

**Table 2.**
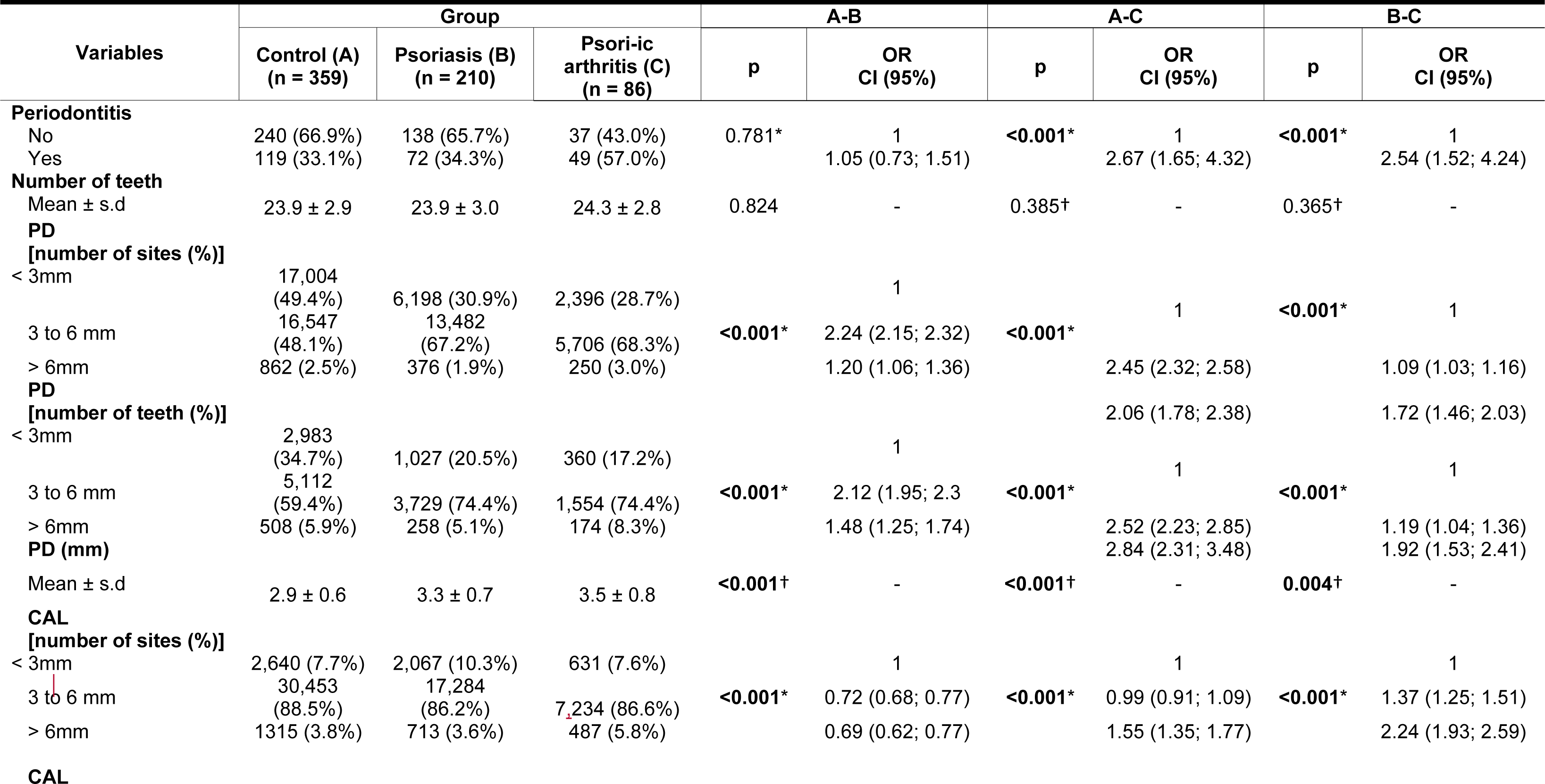
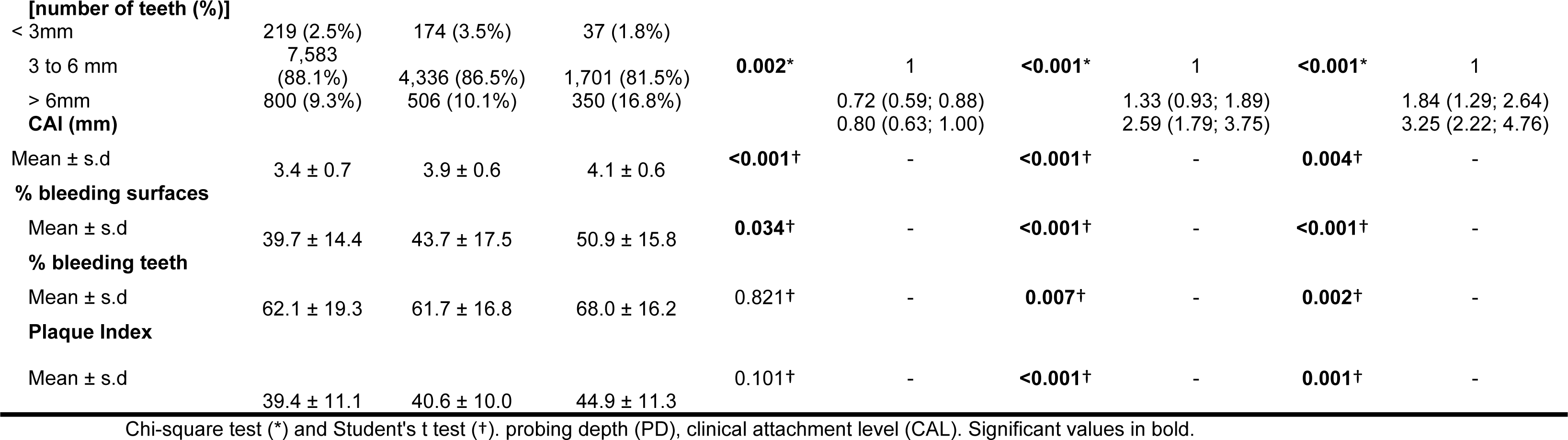
Periodontal parameters of the studied groups.

When analyzing OHRQoL, PsA group presented a significantly higher mean impact (p= 0.011) than controls (Supplementary Figure 1). Interestingly, when using the OIDP stratification into no impact (0), low (1–15), medium (16–45) and high (45–200) scores a significant difference was identified among PSO and PsA individuals, who have a higher mean of high OIDP scores when compared to controls (p<0.001). High OIDP scores indicate worse OHRQoL.

Table 3 shows the characterization of individuals regarding each OIDP activity, in frequency and severity. A significant difference exists when there is no overlap between CI as in the comparison of the frequency of discomfort when speaking between patients with and without PE in the control group: those with PE experienced discomfort when speaking more frequently than those without PE. It is observed that in the control and PSO groups, 11 items do not overlap, indicating a significant difference between them, and that they are worse in the presence of PE. In the PsA group, the situation is reversed and 11 items overlap. This result may suggest that PsA disease is more debilitating and has a greater impact on the individual’s overall QoL, so that they feel and report less discomfort in the presence of PE or even worry less about their oral condition.

**Table 3.** Comparison between groups in relation to frequency and severity for each OIDP item.

| OIDP | Control |  | Psoriasis |  | Psoriatic arthritis |  |
| --- | --- | --- | --- | --- | --- | --- |
|  | -PE | +PE | -PE | +PE | -PE | +PE |
| **Frequency** |  |  |  |  |  |  |
| Eating | 0.46 (0.35; 0.58) | 0.47 (0.29; 0.67) | **0.43 (0.30; 0.57)** | **2.10 (1.60; 2.58)** | **0.59 (0.33; 0.86)** | **1.78 (1.16; 2.39)** |
| Speaking | **0.04 (0.01; 0.08)** | **0.22 (0.13; 0.32)** | **0.07 (0.03; 0.12)** | **1.40 (0.97; 1.87)** | **0.05 (0.00; 0.14)** | **0.94 (0.45; 1.43)** |
| Tooth cleaning | 0.30 (0.21; 0.40) | 0.52 (0.34; 0.72) | **0.30 (0.18; 0.43)** | **2.17 (1.72; 2.65)** | **0.32 (0.08; 0.59)** | **1.41 (0.84; 1.98)** |
| Smiling | **0.33 (0.24; 0.43)** | **0.11 (0.03; 0.21)** | 0.10 (0.05; 0.17) | 0.42 (0.17; 0.75) | **0.19 (0.08; 0.32)** | **0.76 (0.33; 1.29)** |
| Sleeping | **0.00 (0.00; 0.00)** | **0.22 (0.11; 0.35)** | **0.02 (0.10; 0.33)** | **0.72 (0.39; 1.11)** | 0.27 (0.05; 0.51) | 0.65 (0.24; 1.06) |
| Maintaining emotional state | **0.01 (0.00; 0.04)** | **0.34 (0.18; 0.53)** | **0.19 (0.09; 0.30)** | **0.67 (0.31; 1.04)** | 0.16 (0.00; 0.38) | 0.18 (0.00; 0.47) |
| Activities | **0.01 (0.00; 0.03)** | **0.08 (0.03; 0.13)** | 0.09 (0.03; 0.17) | 0.25 (0.07; 0.49) | 0.30 (0.05; 0.57) | 0.24 (0.00; 0.57) |
| Leisure | 0.18 (0.12; 0.24) | 0.17 (0.09; 0.25) | **0.23 (0.14; 0.33)** | **0.72 (0.39; 1.12)** | 0.51 (0.27; 0.78) | 0.37 (0.08; 0.73) |
| **Gravity** |  |  |  |  |  |  |
| Eating | 0.57 (0.45; 0.7) | 0.61 (0.43; 0.82) | **0.49 (0.28; 0.70)** | **2.10 (1.63; 2.57)** | 0.86 (0.43; 1.38) | 1.59 (1.04; 2.16) |
| Speaking | **0.03 (0.00; 0.06)** | **0.38 (0.24; 0.54)** | **0.38 (0.24; 0.56)** | **1.40 (0.97; 1.87)** | 0.49 (0.22; 0.81) | 1.04 (0.53; 1.53) |
| Tooth cleaning | **0.20 (0.12; 0.28)** | **0.61 (0.41; 0.81)** | **0.28 (0.16; 0.42)** | **2.03 (1.60; 2.50)** | **0.32 (0.08; 0.65)** | **1.22 (0.73; 1.71)** |
| Smiling | **0.45 (0.34; 0.57)** | **0.17 (0.07; 0.30)** | 0.38 (0.22; 0.54) | 0.44 (0.17; 0.78) | 0.41 (0.14; 0.73) | 0.65 (0.24; 1.14) |
| Sleeping | **0.00 (0.00; 0.00)** | **0.25 (0.12; 0.39)** | **0.09 (0.02; 0.17)** | **0.69 (0.36; 1.07)** | 0.24 (0.03; 0.49) | 0.65 (0.24; 1.06) |
| Maintaining emotional state | **0.03 (0.00; 0.06)** | **0.42 (0.26; 0.60)** | 0.20 (0.10; 0.33) | 0.61 (0.28; 0.94) | 0.35 (0.11; 0.65) | 0.16 (0.00; 0.41) |
| Activities | **0.01 (0.00; 0.03)** | **0.14 (0.07; 0.24)** | 0.17 (0.07; 0.30) | 0.33 (0.11; 0.61) | 0.11 (0.00; 0.27) | 0.24 (0.00; 0.57) |
| Leisure | 0.30 (0.19; 0.40) | 0.37 (0.22; 0.52) | **0.17 (0.06; 0.30)** | **0.93 (0.56; 1.38)** | 0.24 (0.05; 0.49) | 0.78 (0.33; 1.22) |

\*Bootstrap range. The values marked in bold indicate the non-overlapping CIs comparing the groups with and without periodontal disease, that is, those with significant differences.

When evaluating the influence of variables of interest on QoL in a dichotomous manner (without/with impact), Table 4 shows that alcohol and anxiolytics use, presence of periodontitis, and psoriasis with periodontitis were significant (p<0.001). In the final logistic regression model, PE (p<0.001) and alcohol/anxiolytic use (p=0.002) remained significantly (Table 5).

**Table 4.**
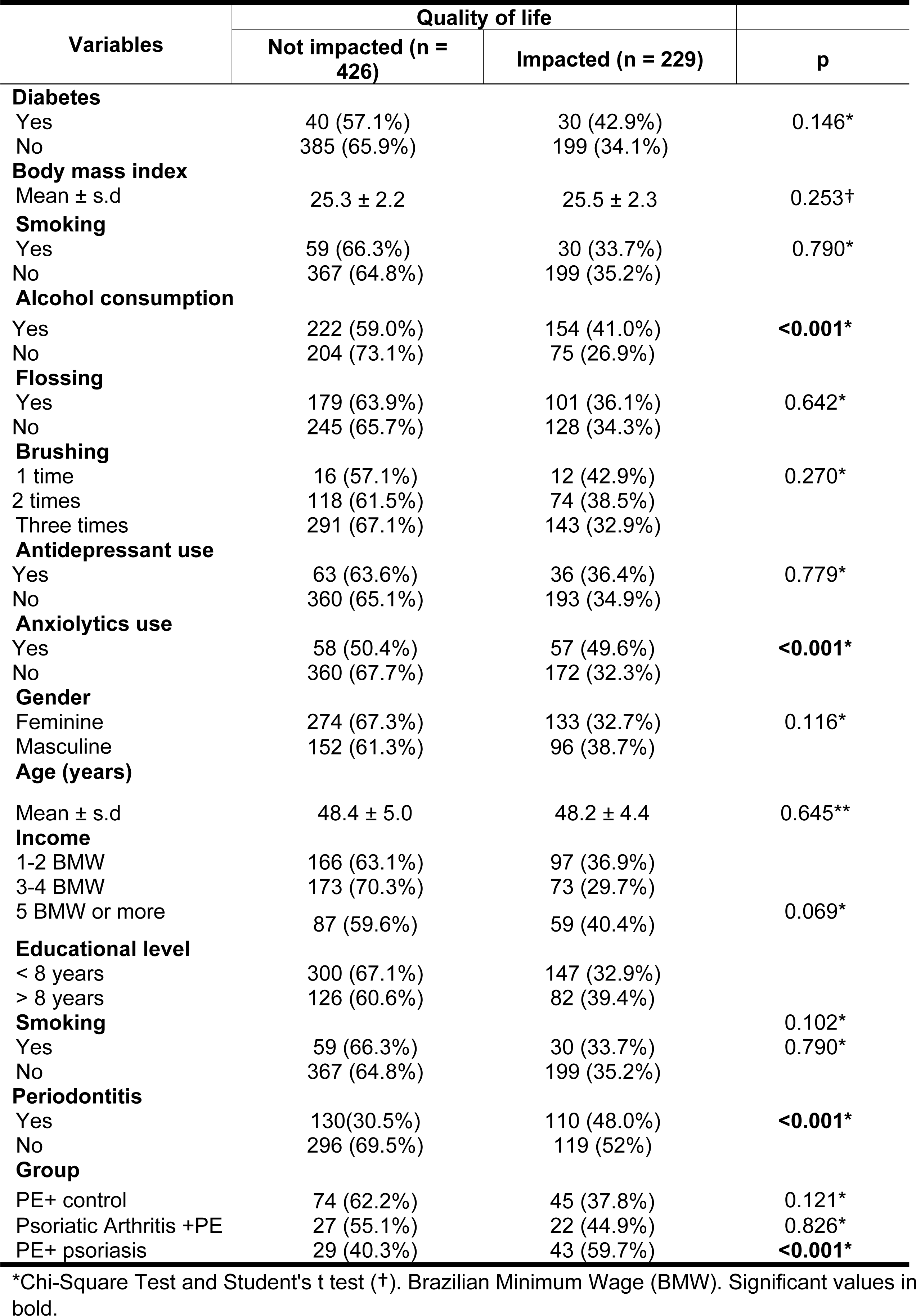
Dichotomous influence of variables of interest on quality of life.

**Table 5.** Logistic regression of variables of interest associated with impacts on quality of life.

| Variables | Full model |  |  | Final model |  |  |
| --- | --- | --- | --- | --- | --- | --- |
| | $\beta$ | p | OR (95% CI) | $\beta$ | p | OR (95% CI) |
| **Gender (Ref. Female)** |  |  |  |  |  |  |
| Masculine | 0.21 | 0.247 | 1.23 (0.87; 1.74) |  |  |  |
| **Income (Ref. 3 to 4 BMW)** |  |  |  |  |  |  |
| 1 to 2 BMW | 0.24 | 0.222 | 1.28 (0.86; 1.89) | 0.27 | 0.164 | 1.32 (0.89; 1.94) |
| 5 BMW or more | 0.58 | **0.011** | 1.78 (1.14; 2.80) | 0.57 | **0.011** | 1.78 (1.14; 2.77) |
| **Educational level (ref. < 8 years)** |  |  |  |  |  |  |
| > 8 years | 0.39 | **0.038** | 1.47 (1.02; 2.13) |  |  |  |
| **Alcohol consumption (Ref. No)** |  |  |  |  |  |  |
| Yes | 0.53 | **0.004** | 1.69 (1.19; 2.42) | 0.56 | **0.002** | 1.74 (1.23; 2.47) |
| **Anxiolytics use (Ref. No)** |  |  |  |  |  |  |
| Yes | 0.72 | **0.001** | 2.05 (1.32; 3.18) | 0.66 | **0.002** | 1.93 (1.26; 2.95) |
| **Diabetes (Ref. No)** |  |  |  |  |  |  |
| Yes | 0.44 | 0.112 | 1.55 (0.90; 2.66) |  |  |  |
| **Body mass index** | -0.03 | 0.531 | 0.97 (0.90; 1.06) | - | - | - |
| **Group (Ref. Control)** |  |  |  |  |  |  |
| Psoriasis | 0.12 | 0.537 | 1.13 (0.77; 1.67) |  |  |  |
| Psoriatic arthritis | 0.15 | 0.586 | 1.16 (0.68; 1.96) | - | - | - |
| **Periodontitis (Ref. No)** |  |  |  |  |  |  |
| Yes | 0.66 | **< 0.001** | 1.93 (1.34; 2.78) | 0.65 | **< 0.001** | 1.92 (1.36; 2.71) |

Hosmes-Lemeshow test (p = 0.170) / AOC = 0.645 / Sensitivity = 0.245 / Specificity = 0.938 / Pseudo R2 = 9.3%. Brazilian Minimum Wage (BMW). Significant values in bold

Regarding periodontal parameters influence on QoL (Table 6), a significant difference was observed for the presence of PE; PD > 6mm; CAL >6mm (p<0.001); higher plaque index (p=0.002), higher BOP % of sites (p=0.005) and teeth (p=0.006). There was a higher PD<3mm (41.6%) percentage of sites in individuals without impact and sites with PD>6mm in individuals with impact (3.1%) (p<0.001). Also, higher PD>6mm % of teeth (8.1% - p<0.001) in individuals with impact, with the same occurring for CAL > 6mm [% of sites (8.4%) and teeth (14.5%) - (p <0.001)]. These data demonstrated that a worse clinical periodontal condition had a negative impact on the analyzed individuals’ OHRQo (Table 6).

**Table 6.** Association analysis between quality of life and periodontal parameters.

| Variables | Quality of life |  | p |
| --- | --- | --- | --- |
|  | Not impacted<br>(n = 426) | Impacted<br>(n = 229) |  |
| **Periodontitis** |  |  |  |
| Yes | 130 (54.2%) | 110 (45.8%) | **< 0.001\*** |
| No | 296 (71.3%) | 119 (28.7%) |  |
| **Number of teeth** |  |  |  |
| Mean ± s.d | 23.9 ± 3.0 | 24.1 ± 2.7 | 0.558<sup>†</sup> |
| **PD [number of sites (%)]** |  |  |  |
| < 3mm | **16,967 (41.6%)** | 8,631 (39.1%) | **< 0.001\*** |
| 3 to 6 mm | 22,959 (56.4%) | 12,776 (57.8%) |  |
| > 6mm | 803 (2.0%) | **685 (3.1%)** |  |
| **PD [number of teeth (%)]** |  |  |  |
| < 3mm | 2,932 (28.8%) | 1,438 (26.0%) | **< 0.001\*** |
| 3 to 6 mm | 6,755 (66.3%) | 3,640 (65.9%) |  |
| > 6mm | 495 (4.9%) | **445 (8.1%)** |  |
| **PD (mm)** |  |  |  |
| Mean ± s.d | 3.0 ± 0.6 | 3.2 ± 0.8 | **< 0.001<sup>†</sup>** |
| **CAL [number of sites (%)]** |  |  |  |
| < 3mm | 3,626 (8.9%) | 276 (2.7%) | **< 0.001 \*** |
| 3 to 6 mm | 35,785 (87.8%) | 9,050 (88.9%) |  |
| > 6mm | 1,325 (3.3%) | **858 (8.4%)** |  |
| **CAL [number of teeth (%)]** |  |  |  |
| < 3mm | 276 (2.7%) | 154 (2.8%) | **< 0.001\*** |
| 3 to 6 mm | 9,050 (88.9%) | 4,570 (82.8%) |  |
| > 6mm | 858 (8.4%) | **798 (14.5%)** |  |
| **CAL (mm)** |  |  |  |
| Mean ± s.d | 3.6 ± 0.7 | 3.8 ± 0.8 | **< 0.001<sup>†</sup>** |
| **% bleeding surfaces** |  |  |  |
| Mean ± s.d | 41.2 ± 15.7 | 44.8 ± 16.6 | **0.005<sup>†</sup>** |
| **% bleeding teeth** |  |  |  |
| Mean ± s.d | 61.4 ± 18.3 | 65.3 ± 17.9 | **0.006<sup>†</sup>** |
| **Plaque index** |  |  |  |
| Mean ± s.d | 39.7 ± 10.7 | 41.9 ± 11.2 | **0.002<sup>†</sup>** |

Chi-square test (\*) and Mann-Whitney test (<sup>†</sup>); probing depth (PD); clinical attachment level (CAL). Significant values in bold.

## Discussion

The present study demonstrated a risk association between PSO and PE, with PsA individuals having significantly worse OHRQoL indicators. A significantly higher PE prevalence was observed among PsA individuals (57%) when compared to controls (33.1%). In PSO (34.35%), the comparison was not significant despite the higher prevalence. PsA individuals had a 2.67 greater chance of developing PE than controls (OR =2.67; p<0.001). Additionally, they presented a significantly worse periodontal clinical condition, expressed by higher means of PI, BOP, PD, CAL as well as % of sites with PD 3-6 mm and >6 mm.

A high prevalence of PE in psoriatic disease individuals is reported in several studies,(12–14,19,28–30) most of which addressed PSO. The literature does not always categorize PSO and PsA conditions separately. Most studies address PSO and some encompass PsA, (12–14,18) referring to these two categories of disease uniquely in their discussion, using terms such as “psoriatic disease” or even “psoriasis group”. This fact directly impacts the possibility of better understanding the prevalence of PE, specifically in PsA. However, within the available literature, when separately evaluating PSO and PsA, the high prevalence of PE in our study (57%) in PsA is very similar to the literature, as reported in the recent study by Mishra *et al*. (13) [(PsA (58.18%) vs. control (42.72 – p=0.042)]. Due to this gap in the literature, it is of high relevance that new studies should aim at individuals with PsA to better elucidate whether their possible greater immune vulnerability puts them at greater risk for PE, credited to the fact of greater severity of psoriatic disease.

Regarding the worse periodontal condition of PSO individuals, our findings corroborate other recent and robust studies reported (12,14,19,28). Our definition of “robust” was based on full-mouth examinations, significant samples and well-defined criteria for PE and psoriatic disease, bringing greater reliability to the findings.

A possible association between PE and increased risk of immune-mediated inflammatory diseases has been increasingly recognized. Although the biological mechanism that explains the increased risk of psoriatic disease among individuals with PE is unknown, several hypotheses have been proposed. One is related to its shared pathology, since exaggerated immune responses to the microbiota present on the epithelial surface are observed in both conditions, suggesting a genetic predisposition that affects dendritic cells and the expression of toll-like receptors (9,16,31,32). Therefore, these pathologies (PE, PsA and PSO) are immune-mediated and present similar etiopathogenic pathways, involving greater susceptibility of an individual with an immune response deficiency and common risk factors.

Observational studies have also confirmed findings of the association between PE and psoriatic disease (12,18,19). However, it is worth mentioning some limitations of these studies, such as: (i) use of retrospective data obtained from medical records(12,14); (ii) use of periodontal indexes to define PE(18,28,29); (iii) small samples(18,28,29); (iv) self-reported information on periodontal conditions and PSO(29,33) and (v) causality bias, as the diseases share numerous risk factors (e.g. smoking and diabetes) (6,15). To date, these limitations may together be an important factor in the presence of conflicting results and moderate evidence for an association between these diseases.

This study demonstrated that individuals with PSO and PsA showed significantly higher BMI values, anxiolytics/antidepressants use and occurrence of diabetes than controls. Those with PSO had greater alcohol consumption than controls (p=0.001), and those with PsA had greater alcohol (p=0.040) and antidepressants (p=0.036) use than PSO individuals. These data corroborate previous findings, since these variables can contribute to systemic triggers activation leading to immunological changes (exacerbation in the expression of cytokines), endocrine and behavioral disorders (lack of hygiene, smoking and other harmful habits) which can predispose greater susceptibility to these diseases (4).

It is noteworthy that mood disorders (particularly depression) have been suggested to be significantly more prevalent in PSO individuals (up to 62%) than in the general population (4 to 10%)(10). Other mental health comorbidities such as anxiety and suicidal ideation have been reported to be more prevalent in psoriatic disease individuals (34).

Contributing factors for this higher prevalence have been reported as: (1) the disease physical impact, with compromised dermal integrity and symptoms such as pain, itching, joint stiffness (35); (2) the clinical signs of the disease, such as skin disfigurement, negatively and directly impacting self-perception of one’s own body (35) and (3) psychiatric disorders that may result in and contribute to the progression of psoriatic disease, suggesting that psoriatic disease and psychiatric conditions (e.g. depression) may have overlapping biological mechanisms – explained by an inflammatory model (10,34).

Several studies have reported that PSO strongly influences global QoL of psoriatic individuals(36–38) and a recent systematic review indicates a higher prevalence of depression and anxiety in PsA, demonstrating worse QoL of these individuals(39).

As evidence suggests that psoriatic patients have worse oral conditions and worse periodontal parameters, it can be assumed that the QoL of these individuals is even more negatively affected by oral changes. Literature already corroborates to the fact that oral conditions such as pain, cavities, tooth mobility and tooth loss, have negative effects on the well-being of individuals, especially in those with PE (38,40).

However, this is the first study that addresses OHRQoL in PsA individuals to date, particularly targeting periodontal condition. Only one recent study(41) evaluated OHRQoL in PSO. The authors reported a risk association between PSO and PE, with individuals with both diseases having significantly worse OHRQoL indicators. In addition, PE and PSO severity significantly increased these negative effects.

The hypothesis proposed in the study that individuals with PsA and PE would present worse OHRQoL indicators was confirmed. Results demonstrated that PsA group had significantly higher mean impact on OHRQoL (p= 0.011) than the controls. A significant difference was identified among PsA and PSO individuals, who had higher means of high OIDP scores when compared to controls (p<0.001). High OIDP scores indicate worse QoL. These data reinforce the existing, albeit scarce, literature on the impact of psoriatic disease on OHRQoL (38,41).

A higher percentage of individuals with an impact on QoL was observed in those with PE (45.8%) when compared to those without PE (28.7%) when evaluating QoL and periodontal parameters Thus, our findings corroborate those of negative impacts of PE on OHRQoL, reinforcing the literature that points out that worse periodontal conditions negatively and significantly affect individuals’ routine activities. Literature reports studies using the OIDP to assess OHRQoL (40–43) while others used different instruments, such as the Oral Health Impact Profile questionnaire (OHIP-14) (44,45).

Some limitations must be attributed to the present study. The main one being inherent to a case-control study which does not demonstrate a causal link due to temporality; with our data, it is not possible to determine whether PE led to greater susceptibility to psoriatic disease or whether individuals with psoriatic disease develop more PE. Patient-reported outcomes (PROMs) such as the questionnaire application in an interview form may have potential interviewer bias, however the extensive impact grading [from 0 (null) to 5 (maximum)] tends to minimize this effect.

On the other hand, advantages can also be cited as (i) high number of individuals in the sample, increasing the statistical power of the study, since PSO has a low prevalence, making it difficult to obtain large samples; (ii) PSO and PsA diagnosis by specialized physicians; (iii) full mouth periodontal examination with robust criteria for PE definition, as it is recognized that the quality of periodontal data and the criteria for defining PE strongly impacts the results of association studies(46); and (iv) uniform methodology for clinical assessment and questionnaire application. However, we highlight the need for additional studies in different populations, with different study designs to expand our knowledge on OHRQoL in psoriatic disease patients.

Thus, our study can be considered an important starting point for investigations into OHRQoL PsA patients and the association between PE and psoriatic disease, highlighting the inclusion of PE as another important comorbidity associated mainly with PsA. Also, the need for attention to the oral condition and its impacts of OHRQoL by both dentists and doctors of these individuals who already present greater physical-emotional vulnerability is advised.

Periodontists and dermatologists must broaden their clinical vision, examining the oral cavity and dermatological lesions, providing a holistic view of the individual. Multidisciplinary interaction is desirable to improve the impact of these diseases on the QoL of individuals with psoriatic disease and PE.

## Conclusion

This study demonstrated the negative impacts of PE on the OHRQoL of the analyzed individuals, being PSO, mainly PsA and PE individuals presenting significantly worse OHRQoL indicators.

## Data Availability

The data underlying the results presented in the study are available in the University's Postgraduate Department Universidade Federal de Minas Gerais. Contact information: https://www.odonto.ufmg.br/posgrad/

## Acknowledgments

This study was supported by the National Council for Scientific and Technological Development - CNPq, Brazil (Productivity research grants n° 307034/2015-1; n° 307024/2015-6 and funding n° 402158/2016-4).

---

**Supporting Figure 1.**
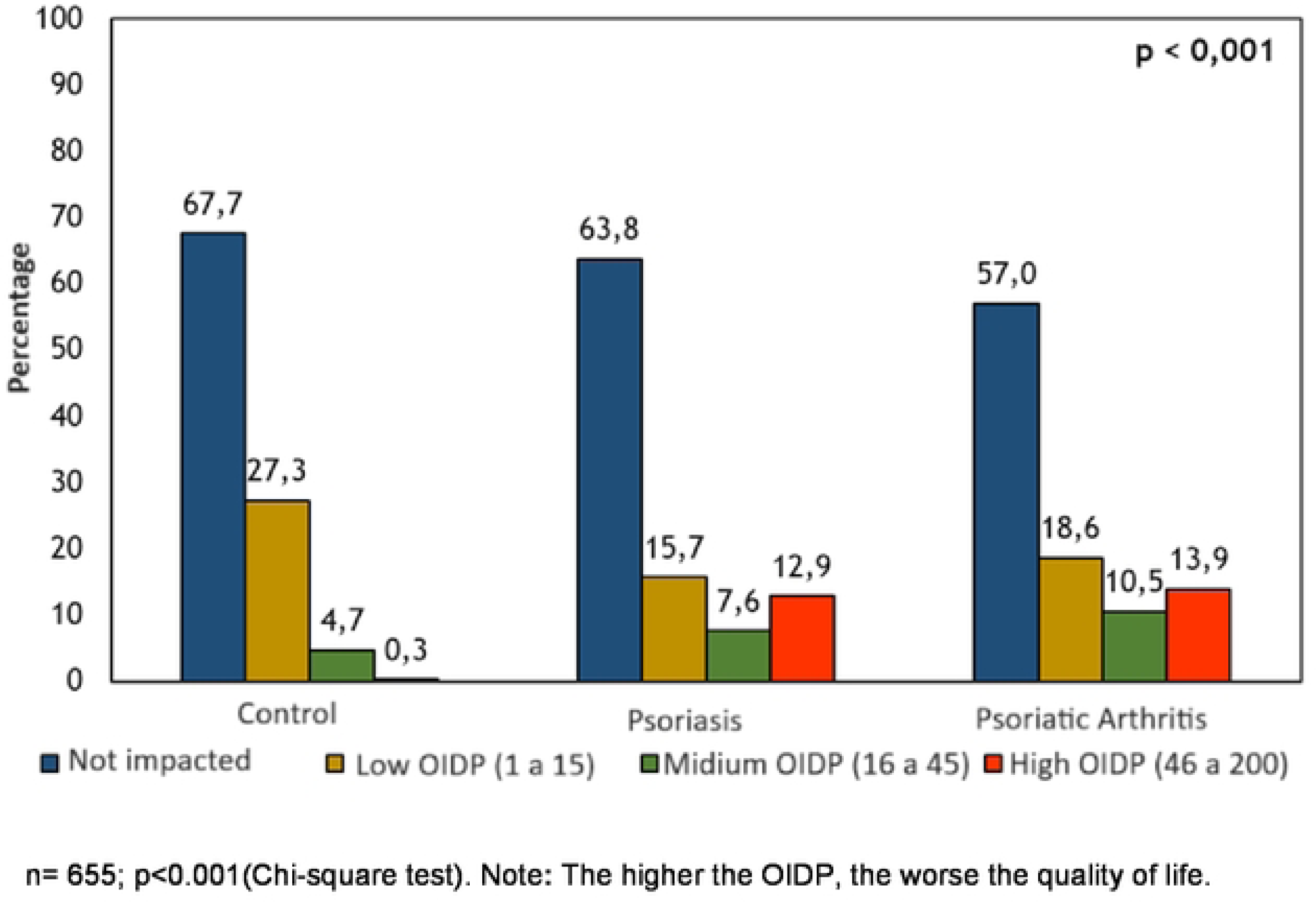
Stratification of OIDP scores between groups.

3 Open Epidemiologic Statistics for Public Health, version 3.01, Boston, MA, USA.

4 UNC-15, Hu-Friedy, Chicago, IL, USA.

5 R software originally created by Ro ss Ihaka and Robert Gentleman, University of Auckland, New Zealand (version 3.6.1).

